# FCFNets: A Factual and Counterfactual Learning Framework for Enhanced Hepatic Fibrosis Prediction in Young Adults with T2D

**DOI:** 10.1101/2025.03.07.25323577

**Authors:** Qiang Yang, Anu Sharma, Daphne Calin, Chloe de Crecy, Rohit Inampudi, Rui Yin

## Abstract

Hepatic fibrosis poses a significant health risk for young adults with type 2 diabetes (T2D). We propose FCFNets, a novel factual and counterfactual learning framework to predict hepatic fibrosis in young adults with T2D that can address class imbalance issue and increase interpretability leveraging electronic health records (EHRs). We designed a hybrid UNDO oversampling strategy, combining random and dissimilar oversampling that improves dataset diversity and model robustness. FCFNets also integrates SHAP-based global and instance-level explanations, alongside feature interaction analysis, providing insights into critical risk factors associated with hepatic fibrosis. The results show our proposed model outperforms various baseline methods with high sensitivity (0.846) and accuracy (0.768), while delivering counterfactual explanations. Hyperparameter tuning and dropout analysis further refine the model, ensuring optimal performance. This study demonstrates FCFNets’s potential for early detection and personalized management of hepatic fibrosis, paving the way for interpretable AI applications in precision medicine.

## Introduction

Metabolic dysfunction-associated liver disease (MASLD), previously referred to as non-alcoholic fatty liver disease (NAFLD), is the presence of steatosis (fatty infiltrates) in the liver in the setting of at least one metabolic disease and without no other known cause of liver disease including heavy alcohol use. Type 2 diabetes mellitus (T2D) markedly increases the risk of MASLD. As such, standard of care guidelines recommend screening for MASLD as part of the comprehensive medical visit^1^. MASLD can progress to steatohepatitis with or without clinically significant hepatic fibrosis (fibrosis stage ≥2; F≥2). This is referred to as “at-risk” metabolic dysfunction-associated steatohepatitis^2^ (MASH). People with “at-risk” MASH have a higher rate of developing cirrhosis, liver failure and death^3,4^. Early identification of “at-risk” MASH is therefore critical to prevent future cirrhosis. To date, liver biopsy has been the gold standard to diagnose and stage MASLD. It is an invasive procedure that requires expertise in both the intervention and interpretation, is costly, and has complications including death^5^. Non-invasive strategies to identify those with “at-risk” MASH include clinical risk scores, laboratory-based biomarkers, and imaging. For example, the fibrosis-4 (FIB-4) test is the most validated clinical risk score to stratify adults based on serum aminotransferase concentrations, platelets, and age^6^. However, the sensitivity of FIB-4 is lower in people with T2D (48.5-79.5%)^7^ and it is much more unreliable in younger adults <35 years old (0-46.7%)^8^. Therefore, accurately identifying “at-risk” MASH in young adults with T2D remains a significant challenge.

The electronic health record (EHR) provides a valuable resource, aggregating information such as demographics, comorbidities, laboratory results, and treatments. Machine learning (ML) methods have been widely employed to leverage EHR data for identifying liver-related diseases^9,10^. For example, Thrift et al. employed logistic regression and random forest classification to predict hepatic fibrosis on 344 participants who underwent Fibroscan examination using EHR data^9^. Ghandian et al. developed a gradient boosted machine learning algorithm (XGBoost) to identify patients with newly diagnosed NAFLD who were at high risk of progression to steatohepatitis or fibrosis^11^. However, these methods face significant challenges due to class imbalance, where patients in young adults with T2D are usually under-diagnosed for the onset of hepatic fibrosis. This issue often results in imbalanced data distribution that may cause biased models disproportionately favoring the majority class, leading to suboptimal performance^12,13^. To address this challenge, various data sampling methods have been developed to mitigate the impact of imbalance, including undersampling, oversampling, synthetic sampling, and hybrid approaches^14,15^.

In this study, we propose FCFNets, an interpretable factual and counterfactual learning framework to predict “at-risk” MASH in young adults with T2D. FCFNets combines two complementary components, namely, factual learning and counterfactual learning. The factual learning focuses on traditional supervised learning for predictions, while the counterfactual learning generates hypothetical scenarios to examine how the changes of input features could alter predictions. Specifically, a shared layer first encodes the input features into a latent status. Then, a multi-layer perceptron (MLP) is employed as the predictor for factual learning, while a decoder that incorporates the desired class facilitates counterfactual learning. This integrative approach could not only enhance prediction accuracy but provide insights into the factors driving progression to “at-risk” MASH in young adults with T2D. In addition, we introduce an unbiased and diversity-aware oversampling strategy, called UNDO, which combines random and dissimilar sampling to balance the dataset while maintaining minority class diversity. Specifically, random sampling ensures that all minority samples are equally represented, while dissimilar sampling prioritizes samples least like the majority class, enhancing the diversity and characteristics of the minority group. FCFNets also leverages SHAP-based global explanations, instance-level factual and counterfactual interpretation, and feature interaction analysis to uncover the underlying drivers for “at-risk” MASH. The overall workflow of our proposed framework is illustrated in **Figure 1**. We demonstrate the effectiveness of FCFNets on a real-world dataset from the University of Florida (UF) Health Integrated Data Repository (IDR)^1^. Extensive experiments suggest the superior performance of FCFNets compared to different benchmarks, including traditional machine learning models, deep learning methods, and non-invasive diagnostic measurements. We also evaluate the impact of the UNDO oversampling strategy on addressing class imbalance, showing that it significantly improved the predictive performance and robustness of FCFNets.

**Figure 1:**
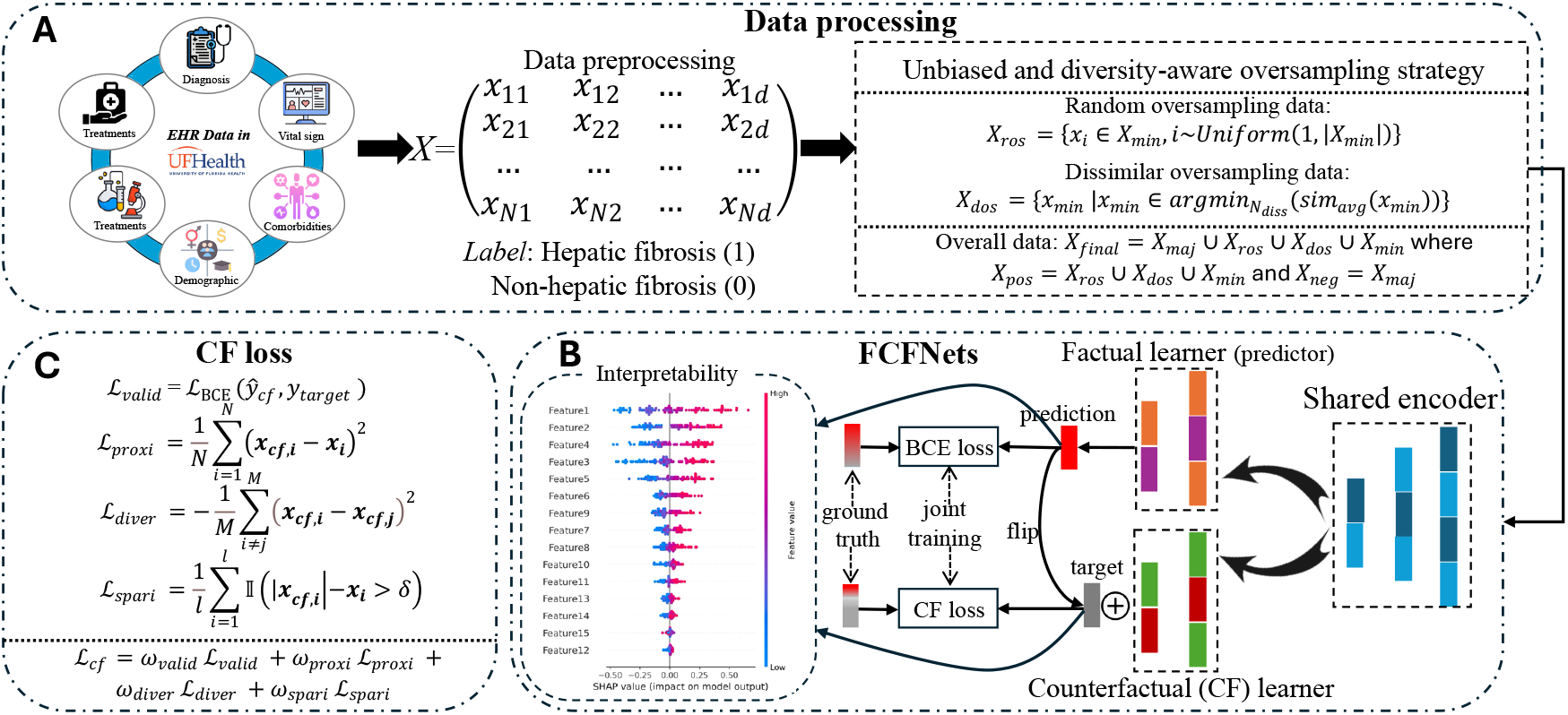
Illustration of the proposed framework. (A) Data processing: the study participants from UFHealth and patient feature selection. (B) Framework of the factual and counterfactual learning with interpretation for identifying hepatic fibrosis in young adults with T2D. (C) The design of counterfactual loss.

## Materials and Methods

### Data source and participant selection

The dataset for this study was obtained from the University of Florida (UF) Health Integrated Data Repository (IDR), a secure clinical data warehouse developed under the UF Clinical and Translational Science Institute and UF Health Information Technology programs. The IDR aggregates data from various UF Health clinical and administrative systems, including the Epic EHR, and currently contains over 1 billion observational facts pertaining to more than 2 million patients. This comprehensive repository includes diverse data types, such as demographics, clinical encounters, diagnoses, procedures, laboratory results, medications, and comorbidity measures. For this study, the cohort was selected by the following inclusion criteria: (1) a diagnosis of type 2 diabetes using International Classification of Diseases (ICD) codes (ICD-9: 250.xx; ICD-10: E11.x), (2) a diagnosis with MASLD using ICD codes (ICD-9: 571.8, 571.9; ICD-10: K74.0, K74.00, K74.01, K74.02, K74.2, K74.6, K75.81, K76.0), (3) age between 18 and <45 years old at the first diagnosis of T2D, and (4) chart review by experts. Exclusion criteria were: (1) no history of T2D or diagnosis of other types of diabetes (e.g. type 1 diabetes, monogenic diabetes, cystic fibrosis etc.), (2) age ≥45 years at the first diagnosis of T2D, (3) pregnancy at the time of diagnosis of liver disease, and (4) other causes of liver disease e.g. viral hepatitis, Wilson’s disease, autoimmune hepatitis, amyloidosis, sarcoidosis, hemochromatosis, alcoholic liver disease etc. In total, 412 cases were identified, comprising 62 patients with “at-risk” MASH (ground truth diagnosis in clinics) and 350 control cases (diagnosed MASLD without fibrosis). The study has been approved by the University of Florida Institutional Review Board (protocol no. IRB202300939).

### Feature preprocess and selection

The clinical information of selected patients consisted of diverse and numerous features that can be categorized into continuous (e.g., body mass index, age) and categorical features (e.g., sex, diagnostic codes). Continuous features were normalized using Min-Max scaling to map values to a 0–1 range, ensuring comparability across features, while categorical features were transformed through one-hot encoding. To preserve data quality, features with a missingness rate exceeding 30% were excluded. A robust pipeline of feature selection techniques was then applied to eliminate irrelevant variables, including (1) the use of a variance threshold to remove features with variance below 0.1, (2) Pearson correlation to retain features with a correlation coefficient greater than 0.01 with the target variable, (3) Chi-Squared tests (p-value < 0.05), and (4) ANOVA tests (p-value < 0.05). We finally obtained 26 distinct clinical features for the subsequent diagnosis of hepatic fibrosis of young adult patients with T2D. The missing values in these selected features were imputed using K-nearest neighbors (k = 3) algorithm to ensure completeness for model training.

### Unbiased and diversity-aware data oversampling strategy

To address class imbalance issue in the cohort that may potentially impair our model, we proposed an unbiased and diversity-aware data oversampling strategy (UNDO). It can effectively preserve the diversity and representativeness of the minority class by combining random oversampling with dissimilar oversampling using a combination ratio *cr* (0 ≤ *cr* ≤ 1). On one hand, random oversampling duplicates existing minority class samples to ensure unbiased features, ensuring each instance an equal chance of selection; on the other hand, dissimilar oversampling generates diverse and semantically meaningful samples by emphasizing those that are not like the majority class, thereby enhancing the model’s ability to distinguish between classes. We assume *X*_*min*_ represents the set of minority class samples and *N*_*rand*_ denotes the desired number of samples needed for balance. The samples generated by random oversampling are defined as:

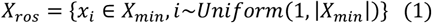

where samples *x*_*i*_ are drawn randomly with replacement until |*X*_*min*_| = *N*_*rand*_. In dissimilar oversampling, we focus on duplicating minority class samples that are not like the majority class samples, ensuring that diverse and distinct characteristics of the minority class are represented. For this, pairwise cosine similarity between minority (*X*_*min*_) and majority (*X*_*maj*_) class samples is computed as:

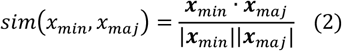

where *x*_*min*_ ∈ *X*_*min*_ and *x*_*maj*_ ∈ *X*_*maj*_ are the minority and majority data samples, respectively. ***x***_*min*_ and ***x***_*maj*_ are the corresponding features. For each *x*_*min*_, the average cosine similarity to all *X*_*maj*_ is calculated:

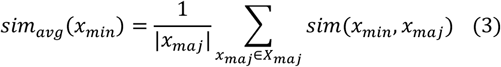

The minority samples with the lowest *sim*_*avg*_ values are identified as the least similar and prioritized for duplication:

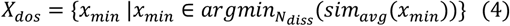

where *N*_*diss*_ is the number of least similar samples selected for oversampling. In addition, *N*_*rand*_ and *N*_*diss*_ satisfy *N*_*rand*_: *N*_*diss*_ = *cr*. The final oversampled dataset combines the samples from both random oversampling and dissimilar oversampling: *X*_*final*_ = *X*_*maj*_ ∪ *X*_*ros*_ ∪ *X*_*dos*_ ∪ *X*_*min*_. Furthermore, the positive-to-negative data ratio, denoted as *pnr*, is defined by *X*_*pos*_: *X*_*neg*_ = *pnr*, where *X*_*pos*_ = *X*_*ros*_ ∪ *X*_*dos*_ ∪ *X*_*min*_ and *X*_*neg*_ = *X*_*maj*_.

### Factual and counterfactual learning

Drawing on the principles of factual and counterfactual reasoning detailed in existing works^16–18^, we developed Factual and Counterfactual Learning Networks (FCFNets) that integrates both learning paradigms. In this framework, factual learning captures the underlying patterns in the observed data, while counterfactual learning explores alternative scenarios to enhance model robustness and interpretability. The architecture comprises three main components: a shared encoder, a factual learner, and a counterfactual learner.

#### Shared Encoder for Features Learning

Given the input data *x* with the features ***x*** ∈ ℝ^*d*^ (where *d* is the input feature dimension), the shared encoder maps ***x*** into a latent feature ***z*** ∈ ℝ^*h*^ : ***z*** = *f*_*enc*_(***x*;** *θ*_*enc*_) (*h* is the latent status dimension). Here, *f*_*enc*_ is implemented as a two-layer multi-layer perceptron (MLP), and *θ*_*enc*_ is the parameter in *f*_*enc*_. It is optimized to produce a latent feature ***z*** that captures meaningful features relevant for both factual prediction and counterfactual generation. The encoder output ***z*** is then fed into both the factual learner and counterfactual learner.

#### Factual Learner

The factual learner predicts the outcome *ŷ* ∈ [0,1] from the latent features ***z***. Formally, it is denoted as: *ŷ* = *f*_*fact*_(***z*;** *θ*_*fact*_) where *f*_*fact*_ is a two-layer MLP incorporating batch normalization and dropout for regularization, and *θ*_*fact*_ is the parameter in *f*_*fact*_. Specifically, the predicted outcome *ŷ* is computed via the sigmoid activation function: *ŷ* = *σ*(*W*_1_ · :*Relu*(*W*_1_***z*** + *b*_1_); + *b*_1_) where *W*_1_, *W*_2_ and *b*_1_, *b*_2_ are learnable parameters of the factual learner. The objective function is defined as the binary cross-entropy loss between *ŷ* and the ground truth *y*, where *N* is the number of samples.

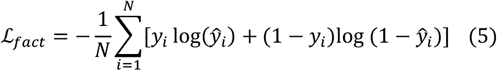

#### Counterfactual Learner

The counterfactual learner generates counterfactual samples *x*_*cf*_ by conditioning on the latent features ***z*** and a specified target class *y*_*target*_. The counterfactual generation is modeled as: ***x***_***cf***_ = *f*_*cf*_ (***z***, *y*_*target*_; *θ*_*cf*_) where *f*_*cf*_ is a two-layer MLP and *θ*_*cf*_ is the parameter in *f*_*cf*_. This module is designed to reconstruct the input features ***x*** while aligning the generated sample to the target class *y*_*target*_. To achieve these goals, the counterfactual loss *ℒ*_*cf*_ is formulated as four parts including validity loss, proximity loss, diversity loss, and sparsity loss. (1) Validity loss ensures that the counterfactual examples achieve the desired target outcomes by penalizing the difference between the predicted counterfactual labels (*ŷ*_*cf*_) and the specified target labels (*y*_*target*_): *ℒ*_*valid*_ = *ℒ*_BCE_(*ŷ*_*cf*_, *y*_*target*_), where *ℒ*_BCE_ represents the binary cross-entropy loss. (2) Proximity loss encourages the counterfactual examples (*x*_*cf*_) to remain close to the original input data (*x*) by penalizing large deviations using the mean squared deviation: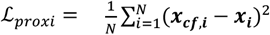, where *N* is the number of all samples. (3) Diversity loss promotes variety among the generated counterfactuals by minimizing the negative average pairwise Euclidean distance, thus preventing the model from producing overly similar samples: 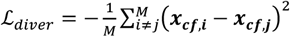, where *M* is the number of counterfactual samples. This loss is clamped to a minimum value to prevent extreme gradients. (4) Sparsity loss encourages minimal yet meaningful changes by penalizing the number of features whose differences exceed a threshold: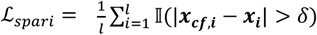, where *l* is total number of features in the input ***x***, and 𝕀(·) is the indicator function. *δ* is a predefined threshold value that determines the significance of changes. If the absolute difference between ***x***_***cf***,***i***_ and ***x***_***i***_ exceeds *δ*, the change is considered significant. The total counterfactual loss is a weighted sum of these components:

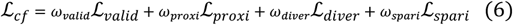

where *ω*_*valid*_, *ω*_*proxi*_, *ω*_*diver*_, and *ω*_*spari*_ are the respective weights for each loss term. This balanced formulation ensures that the generated counterfactuals are valid, interpretable, diverse, and minimally intrusive.

#### Model Training

The overall loss function combines the factual and counterfactual, i.e., *ℒ*_*fact*_ and *ℒ*_*cf*_ to optimize both the prediction accuracy and the quality of counterfactual explanations. It is defined as follows, where *λ* is the weighting parameter to balance the contribution of *ℒ*_*fact*_ and *ℒ*_*cf*_ (*λ* =1 in our experiments).

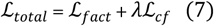

### Model interpretation

Understanding how a model makes predictions is as crucial as its predictive performance, especially in biomedical domains where interpretability fosters trust among patients and clinicians. Accordingly, we employe global explanation, factual and counterfactual interpretations, and feature interaction analysis in FCFNets for model interpretation, which examine the contributions of input features and their interactions in identifying hepatic fibrosis for young T2D adult patients.

#### Global Explanation

A global explanation offers a comprehensive view of the model’s behavior by examining how individual features contribute to predictions across the entire dataset. Using SHAP (SHapley Additive exPlanations) values, we identify the most influential features by calculating their mean absolute contributions, then select the top features for detailed analysis. A summary plot visualizes the impact and distribution of these key features, highlighting their importance in the model’s decision-making process.

#### Factual and Counterfactual Explanations

Factual and counterfactual explanations enhance the interpretability of individual predictions by analyzing feature contributions to both observed outcomes and hypothetical scenarios. For factual explanations, SHAP waterfall plots are generated for randomly selected positive and negative instances, clearly illustrating how specific features drive the model’s predicted label. In contrast, counterfactual explanations examine how feature values would need to change to achieve a different outcome. By generating and visualizing counterfactual SHAP values, this method uncovers actionable insights into the factors that could alter the model’s prediction. Together, these complementary approaches offer a holistic understanding of individual predictions, combining clarity on current decisions with insights into potential alternative outcomes, thereby enhancing interpretability and supporting informed decision-making.

#### Feature Interaction Analysis

This analysis delves into how pairs of features collectively influence model predictions by leveraging SHAP values to quantify and visualize these interactions. For each feature pair, SHAP values are calculated to reveal how one feature’s value affects predictions when conditioned on the other, capturing the joint impact on the model’s output. All pairwise interactions are then evaluated by summing their absolute SHAP contributions to provide a global view of feature interplay, and the top interactions are ranked by their mean interaction strength. This approach can uncover critical insights into how features work in tandem to drive the model’s decisions.

### Experimental setup

#### Implementation and evaluation

The dataset was randomly divided into training (70%), validation (10%), and testing (20%) sets, with hepatic fibrosis labeled as “1” and non-hepatic fibrosis as “0”, representing positive and negative samples, respectively. FCFNets was optimized over 80 training epochs using a batch size of 16 and a learning rate of 0.001. To mitigate overfitting, we implemented a dropout strategy with a rate of 0.3 and employed early stopping, which halted training if performance did not improve for 20 consecutive epochs. The model was trained on the training set and subsequently validated separately on the validation and testing sets. Coefficients in equation 6, including *ω*_*valid*_, *ω*_*proxi*_, *ω*_*diver*_, and *ω*_*spari*_ are set as 0.04, 10.0, 0.03, and 0.05 respectively. To further explore the impact of data oversampling strategies, we experimented with random, dissimilar, similar, and cluster-based oversampling techniques, and performed a sensitivity analysis on key hyperparameters such as oversampling ratios, positive-to-negative ratio, network dimension, and dropout rate to ensure the optimal configuration of our model. We employed five distinct metrics to evaluate the prediction, including sensitivity, specificity, accuracy, F1-score, and the area under the receiver operating characteristic curve (AUROC). For these metrics, higher values indicate better performance. In addition, we assessed the counterfactual learning performance of our model using four metrics: validity, proximity, diversity, and sparsity. Here, higher values are preferable for validity and diversity, while lower values are better for proximity and sparsity.

#### Benchmark approaches

We benchmarked our designed FCFNets against several baseline models across three categories: traditional machine learning (ML) methods, existing deep learning (DL) approaches, and non-invasive clinical measurement for hepatic fibrosis diagnosis. For traditional ML models, we utilized decision tree (DT), random forest (RF), XGBoost (XGB), K-nearest neighbors (KNN), and support vector machine (SVM), each configured with the built-in data balancing techniques available in the Python package scikit-learn. Further, we compared our model with convolutional neural networks^19^ (CNNs), multi-layer perceptron^20^ (MLP), TabNet^21^, autoencoder^22^, and Transformer^23^. We followed the original settings for these benchmarks in the literature. To evaluate the practical utility of our model, we also leveraged several well-known non-invasive measurements in clinics for comparison involving FIB-4^6^, Aspartate Aminotransferase to Platelet Ratio Index (APRI)^24^, and Nonalcoholic Fatty Liver Disease Fibrosis Score (NFS)^25^.

## Results

### Descriptive statistics of patient data

**Table 1** presents the demographic and laboratory characteristics of the selected cohort, consisting of study group (62 patients with “at-risk” MASH) and control group (350 patients without hepatic fibrosis). Overall, patients with “at-risk” MASH are slightly older than those without “at-risk” MASH (40.1 ± 6.5 vs 37.4 ± 7.6 years), though both groups are young adult patients. In terms of sex distribution, female patients outnumbered male patients in both groups. However, the percentage of female patients in each group was relatively close (61.3% in the study group vs. 63.7% in the control group), suggesting that sex may not be a significant indicator of “at-risk” MASH among young adults with T2D. The racial and ethnic breakdown reveals that the study group includes a larger proportion of White patients (74.2% vs 58.6%) and a smaller proportion of Black patients (11.3% vs 27.4%) than the control group. Regarding laboratory measures, the study group exhibits a higher BMI (42.2 ± 9.1 vs 40.4 ± 9.6) on average. Additionally, we found a lower platelet counts (250.5 ± 88.4 vs 273.2 ± 79.3) in patients with “at-risk” MASH, whereas the LDL levels are slightly higher (117.2 ± 31.6 vs. 108.8 ± 38.3). Other laboratory measures, such as AST and ALT, show no substantial differences between the groups.

**Table 1.**
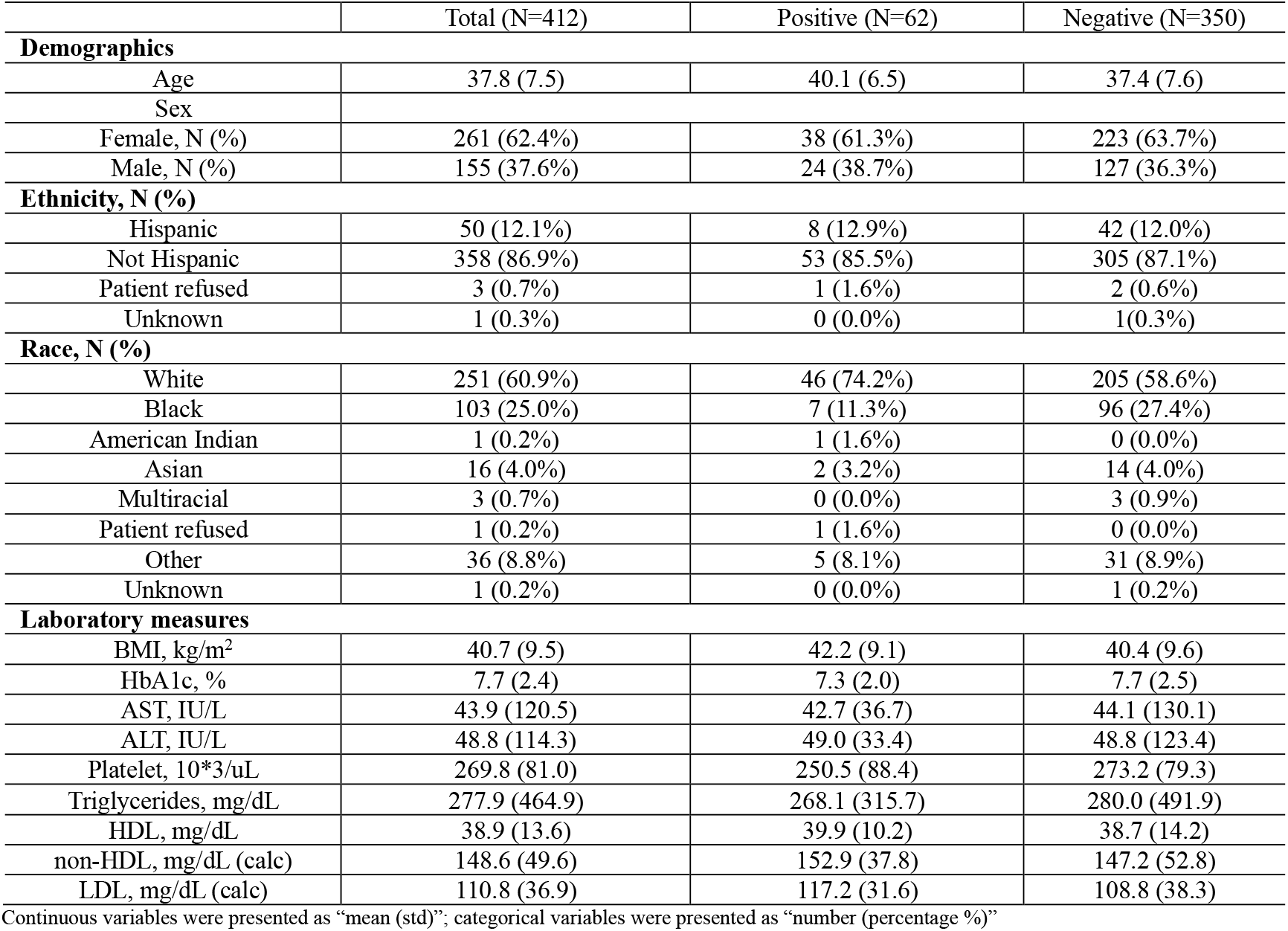
Descriptive statistics on the characteristics of the study cohort.

### Comparative performance of our model against benchmark approaches

We compared the performance of FCFNets against benchmark approaches as shown in **Table 2**. Within deep learning models, autoencoder showed the best performance, achieving the highest accuracy (0.817), along with strong F1-score (0.516) and specificity (0.855), attributable to its ability to learn meaningful latent features. The Transformer model also demonstrated competitive performance by attaining the highest AUROC (0.842). In contrast, despite employing the UNDO data oversampling strategy, both CNN and TabNet underperformed, with low sensitivities (0.231 and 0.462, respectively), indicating a limited ability to leverage oversampled data or handle complex feature interactions effectively. Among traditional machine learning approaches, XGBoost and SVM showed balanced performance, where SVM achieved the highest sensitivity (0.688) and XGBoost suggested a reliable trade-off between sensitivity (0.500) and specificity (0.776). Although KNN excelled in specificity (0.940) and accuracy (0.831), its low sensitivity (0.375) hindered its ability to effectively identify hepatic fibrosis for T2D young patients. Moreover, for the non-invasive clinical measurements, including FIB-4, APRI, and NFS, we found that they exhibited worse performance compared to both ML and DL models. In more detail, FIB-4 demonstrated moderate diagnostic performance with an AUROC of 0.820 with the cutoff 2.67 and 0.712 with the cutoff 1.0, and APRI achieved the best AUROC of 0.848. However, APRI failed to identify any patient with “at-risk” MASH, as reflected by the sensitivity and F1-score of both 0.000. NFS only marginally improved performance with an AUROC of 0.760. One of the most likely explanations is that the traditional clinical non-invasive methods were developed to detect advanced fibrosis in patients with viral hepatitis or in older cohorts^24,25^. As such, they were less sensitive to measure the “at-risk” MASH in young adults. Notably, FCFNets outperformed all baseline models by achieving the highest sensitivity (0.846), a balanced specificity (0.754), and a robust F1-score (0.537). The integration of factual and counterfactual learning, along with the sampling method UNDO, allows it to fully make use of oversampled data with an optimal balance between sensitivity and specificity for “at-risk” MASH prediction.

**Table 2.** Performance comparison between our proposed framework and baseline classifiers. The best and second-best models are marked with bold and underline, respectively. Deep learning methods use the UNDO for data oversampling and machine learning methods use the built-in data balancing strategy. “-” means results are unavailable.

| Benchmark | Models | Sensitivity | Specificity | Accuracy | F1-score | AUROC |
| --- | --- | --- | --- | --- | --- | --- |
| Deep learning approach | MLP | 0.615 | 0.812 | 0.780 | 0.471 | 0.827 |
|  | CNN | 0.231 | 0.866 | 0.762 | 0.240 | 0.502 |
|  | TabNet | 0.462 | 0.594 | 0.573 | 0.255 | 0.590 |
|  | Autoencoder | 0.615 | 0.855 | <u>0.817</u> | <u>0.516</u> | 0.796 |
|  | Transformer | 0.615 | 0.797 | 0.768 | 0.457 | <u>0.842</u> |
| Traditional machine learning classifier | DT | 0.438 | 0.791 | 0.723 | 0.378 | 0.641 |
|  | RF | 0.250 | 0.881 | 0.759 | 0.286 | 0.693 |
|  | XGBoost | 0.500 | 0.776 | 0.723 | 0.410 | 0.738 |
|  | KNN | 0.375 | <u>0.940</u> | **0.831** | 0.462 | 0.763 |
|  | SVM | <u>0.688</u> | 0.701 | 0.699 | 0.468 | 0.778 |
| Non-invasive method | FIB-4 (cutoff=1.00) | 0.407 | 0.767 | - | 0.300 | 0.712 |
|  | FIB-4 (cutoff=2.67) | 0.085 | **0.952** | - | 0.125 | 0.820 |
|  | APRI | 0.000 | 0.848 | - | 0.000 | **0.848** |
|  | NFS | 0.357 | 0.832 | - | 0.312 | 0.760 |
| Our model | FCFNets | **0.846** | 0.754 | 0.768 | **0.537** | 0.818 |

### Factual and counterfactual learning evaluation under different oversampling strategies

To evaluate the influence of different oversampling strategies on FCFNets’ performance, we incorporated additional two methods, i.e., similar oversampling and cluster oversampling, into our analysis. Similar oversampling focuses on augmenting samples most akin to the majority class, in contrast to dissimilar oversampling that generates semantically distinct samples, while cluster oversampling produces additional samples within each cluster of the minority class. For baseline comparison, the default method used the original data without any oversampling. As shown in **Table 3**, the default method achieved perfect sensitivity (1.000) but suffered from extremely low specificity (0.188), preferring to the minority class. Both random and similar oversampling improved overall performance. Our model yielded higher specificity (0.783) and accuracy (0.768) through random oversampling, whereas it showed slightly higher sensitivity (0.769) with similar sampling. When we applied dissimilar oversampling, we could attain the highest sensitivity (0.846) and improved diversity (0.192), underscoring its ability to generate semantically distinct samples. Cluster oversampling suggested limited improvement, with balanced specificity (0.754) but lower sensitivity (0.615) and diversity (0.130). Notably, our proposed UNDO method, which combines the strengths of random and dissimilar oversampling, achieved the best overall balance, with sensitivity and specificity of 0.846 and 0.754, respectively, as well as the highest accuracy (0.768) and F1-score (0.537). For the counterfactual learning evaluation, we found that the validity remained consistently high (1.0) across all methods. However, UNDO stood out by achieving high diversity (0.221) and proximity (0.223) scores, while maintaining low sparsity (8.146). These results highlight UNDO’s effectiveness in generating unbiased and diverse samples that enable both accurate predictions and interpretable counterfactual explanations.

**Table 3.** Performance comparison of factual and counterfactual learning under different oversampling strategies.

| Sampling | Sensitivity | Specificity | Accuracy | F1-score | AUROC | Validity | Proximity | Diversity | Sparsity |
| --- | --- | --- | --- | --- | --- | --- | --- | --- | --- |
| Default | **1.000** | 0.188 | 0.317 | 0.317 | <u>0.822</u> | **1.0** | **0.246** | **0.236** | 10.049 |
| Random | 0.692 | **0.783** | **0.768** | <u>0.486</u> | **0.834** | **1.0** | 0.225 | 0.153 | 8.280 |
| Similar | 0.769 | 0.725 | 0.732 | 0.476 | 0.805 | **1.0** | 0.226 | 0.168 | **7.878** |
| Dissimilar | <u>0.846</u> | 0.681 | 0.707 | 0.478 | 0.818 | **1.0** | 0.224 | 0.192 | <u>8.085</u> |
| Cluster | 0.615 | <u>0.754</u> | <u>0.732</u> | 0.421 | 0.796 | **1.0** | <u>0.232</u> | 0.130 | 8.232 |
| UNDO | <u>0.846</u> | <u>0.754</u> | **0.768** | **0.537** | 0.818 | **1.0** | 0.223 | <u>0.221</u> | 8.146 |

### Model interpretation

We interpreted the predictions made by our FCFNets through a comprehensive visualization approach, as illustrated in **Figure 2**. The global SHAP summary plot (**Figure 2**A) highlights key contributors such as “NITRITE UA_result_negative”, “Abnormal results of liver function studies”, and “Other fatigue”, which consistently show positive SHAP values, underscoring their importance in identifying “at-risk” MASH. Features like “Race”, “Edema, unspecified”, and “Vitamin D deficiency, unspecified” exhibit context-dependent roles, while “Encounter for other preprocedural examination” and “Insomnia, unspecified” show positive associations with fibrosis risk. For a randomly selected positive instance, the factual interpretation (**Figure 2**B) again emphasizes “NITRITE UA_result_negative”, “Abnormal results of liver function studies”, and “Other fatigue” as major contributors, with “Vitamin D deficiency” slightly mitigating the prediction. In the counterfactual interpretation (**Figure 2**C), modifications to features such as “Insomnia, unspecified” and “Edema, unspecified” appear capable of reversing the positive prediction. For a randomly selected negative instance, the factual interpretation (**Figure 2**D) identifies “Race”, “Encounter for other preprocedural examination”, and “Abnormal results of liver function studies” as the strongest factors driving the negative outcome, whereas the counterfactual interpretation (**Figure 2**E) highlights that adjustments in features like “Other nonspecific abnormal finding of lung field” and “Unspecified abdominal pain” could potentially shift the prediction to positive. Finally, the feature interaction analysis (**Figure 2**F) demonstrates robust combined effects, with the top interaction occurring between “NITRITE UA_result_negative” and “Other nonspecific abnormal finding of lung field”. Additional significant interactions involving “Edema, unspecified”, “Deficiency of other specified B group vitamins”, and comorbidities such as “Mixed hyperlipidemia” further emphasize the model’s capacity to capture complex feature relationships, thereby enhancing both interpretability and predictive accuracy.

**Figure 2:**
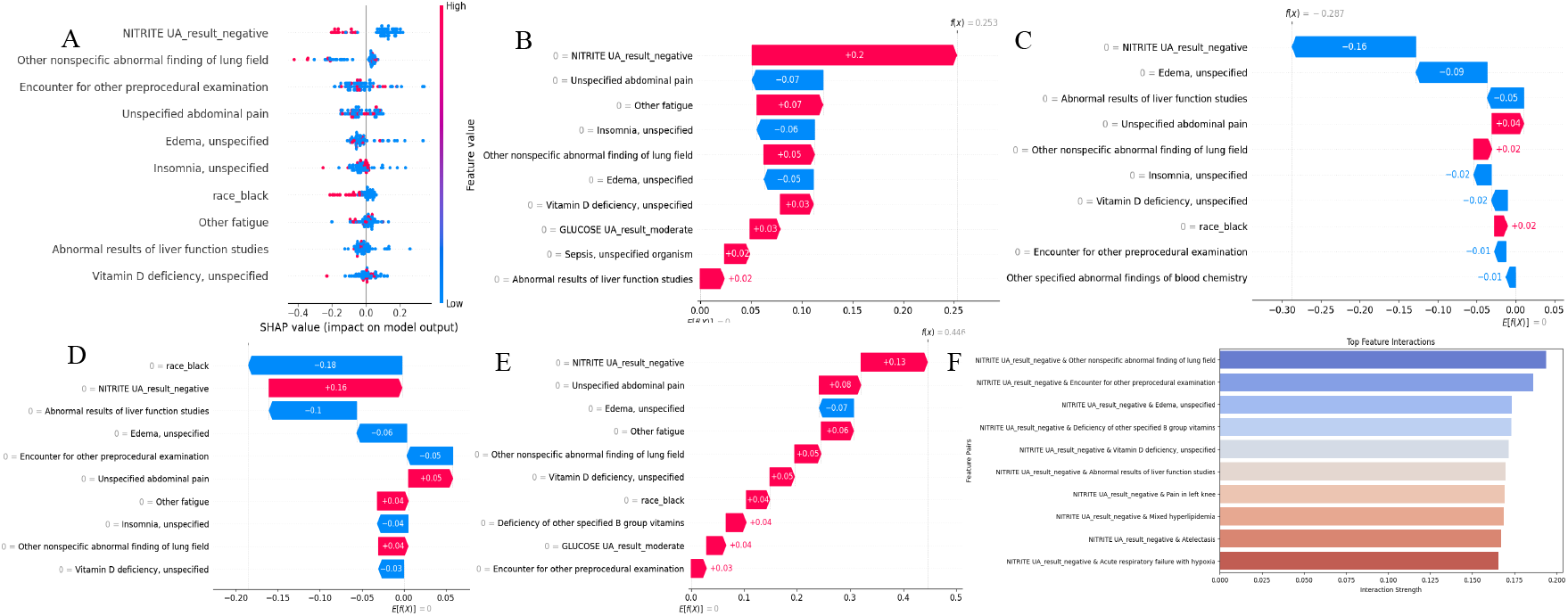
SHAP values of top 10 variables for model’s interpretation. (A) The global interpretation. (B) and (C) are factual and counterfactual interpretation for a randomly selected positive instance, respectively. (D) and (E) are factual and counterfactual interpretation for a randomly selected negative instance. (F) The feature interaction for model’s prediction.

### Parameter analysis

#### Oversampling ratio (cr) in the UNDO method

**Figure 3**A reveals distinct trends in factual and counterfactual metrics across oversampling ratios for FCFNets. When varying the ratio *cr*, i.e., random: dissimilar, the factual metrics exhibit noticeable fluctuations. Notably, at the 8:2 ratio, sensitivity (0.846) and specificity (0.754) are balanced, while accuracy (0.768), AUROC (0.818), and F1 score (0.537) reach a peak. Our counterfactual evaluation shows consistent validity (1.000) across all oversampling ratios, confirming that the generated counterfactuals reliably achieve the desired outcome. Proximity remains stable at 0.223, indicating that counterfactuals are consistently close to the factual instances. Notably, diversity peaks at 0.221 at the 8:2 ratio, suggesting that this setting produces the most distinct counterfactual explanations. While sparsity is lowest at 6:4 (7.317), the 8:2 ratio’s sparsity (8.146) remains acceptable, meaning the extra feature changes do not hinder interpretability. These trends highlight the 8:2 ratio as the best-performing setting, with a strong balance between predictive accuracy and counterfactual interpretability.

**Figure 3:**
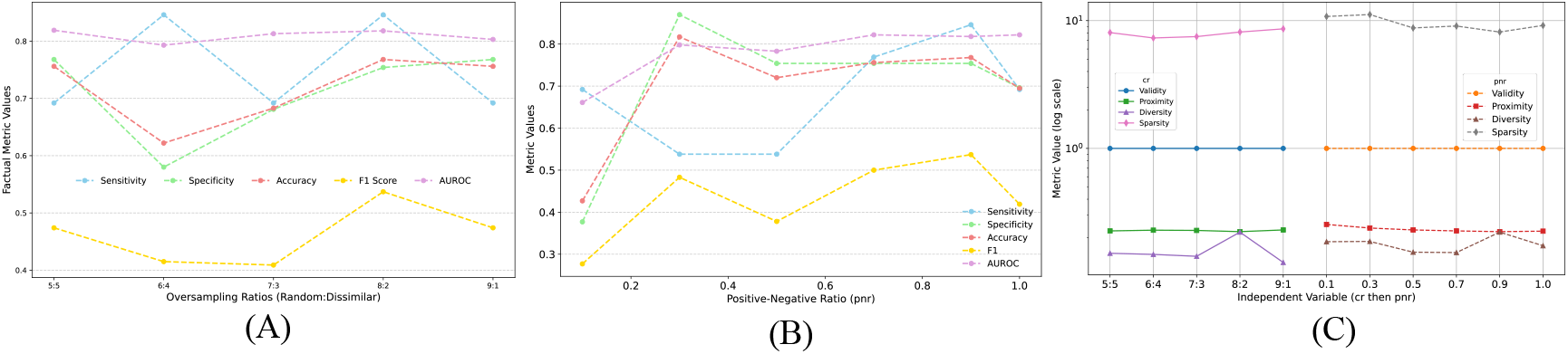
The influence of parameters in UNDO oversampling strategy on FCFNets performance. (A) The oversampling ratio *cr*, i.e., random: dissimilar, affects the performance of factual learning. (B) The positive-to-negative ratio *pnr*, i.e., positive: negative, affects the performance of factual learning. (C) The oversampling ratio *cr* (left hand) and positive-to-negative *pnr* (right hand) affect the performance of counterfactual learning, respectively.

#### Positive-to-negative ratio (pnr) in the UNDO method

**Figure 3**B illustrates the trends in both factual and counterfactual outcomes across different *pnr* in UNDO for FCFNet. Sensitivity reaches a peak at higher *pnr* values (e.g., 0.9), while specificity remains relatively stable at moderate *pnr* levels but declines at extreme values. We achieve the highest accuracy and F1-score at a *pnr* of 0.9. In the counterfactual evaluation, validity holds steady at 1.0 across all *pnr* values, and proximity improves slightly as the *pnr* increases. Moreover, diversity reaches a peak at a *pnr* of 0.9, while sparsity is lowest at this ratio. These trends highlight a *pnr* of 0.9 as the optimal setting for balancing factual performance with counterfactual interpretability.

#### The layers’ dimension in FCFNets

**Table 4** (part A) indicates the influence of layer dimensions on the performance of FCFNet. The increase of the layer dimensions (e.g., from 16/8/4/8 to 64/32/16/32) generally improves performance in most metrics. The 64/32/16/32 configuration achieves the highest sensitivity (0.846), F1-score (0.537), AUROC (0.818) and diversity (0.221), while maintaining balanced proximity (0.223). This demonstrates that larger dimensions effectively capture more complex features. However, extremely large dimensions like 128/64/32/64 result in a slight performance drop in most metrics, such as F1-score (0.474) and increase sparsity (8.878).

**Table 4:** The influence of hyperparameters in the FCFNets. The Part A shows the influence of the layers’ dimension, including *encoder_hidden, encoder_output, pred_hidden*, and *decoder_hidden* on the performance of FCFNet. The part B displays the impact of dropout rate in the factual learner (predictor) on the performance of FCFNet. Sen: sensitivity; Spe: specificity; Acc: accuracy.

| Group |  | Sen | Spe | Acc | F1 | AUROC | Validity | Proximity | Diversity | Sparsity |
| --- | --- | --- | --- | --- | --- | --- | --- | --- | --- | --- |
| Combination<br>(Part A) | 16/8/4/8 | 0.692 | 0.739 | 0.732 | 0.450 | 0.849 | 1.000 | 0.234 | 0.195 | 11.793 |
|  | 32/16/8/16 | 0.538 | 0.870 | 0.817 | 0.483 | 0.794 | 1.000 | 0.249 | 0.138 | 11.439 |
|  | 64/32/16/32 | **0.846** | **0.754** | **0.768** | **0.537** | **0.818** | **1.000** | **0.223** | **0.221** | **8.146** |
|  | 128/64/32/64 | 0.692 | 0.768 | 0.756 | 0.474 | 0.797 | 1.000 | 0.235 | 0.141 | 8.878 |
| Dropout<br>(Part B) | 0.1 | 0.769 | 0.725 | 0.732 | 0.476 | 0.818 | 1.000 | 0.224 | 0.162 | 8.037 |
|  | 0.2 | 0.692 | 0.783 | 0.768 | 0.486 | 0.814 | 1.000 | 0.222 | 0.238 | 7.549 |
|  | 0.3 | **0.846** | **0.754** | **0.768** | **0.537** | **0.818** | **1.000** | **0.223** | **0.221** | **8.146** |
|  | 0.4 | 0.538 | 0.797 | 0.756 | 0.412 | 0.755 | 1.000 | 0.229 | 0.146 | 7.695 |
|  | 0.5 | 0.769 | 0.667 | 0.683 | 0.435 | 0.823 | 1.000 | 0.229 | 0.169 | 7.768 |

#### Dropout rate in FCFNets

**Table 4** (part B) explores the effect of dropout rates on FCFNet’s performance. A moderate dropout rate of 0.3 strikes the optimal balance, yielding the highest sensitivity (0.846), F1-score (0.537), AUROC (0.818), and diversity (0.221). These results prove the role of regularization in preventing overfitting while preserving robust predictions. Although lower dropout rates (0.1 and 0.2) produce comparable results, they slightly underperform in sensitivity and diversity. Conversely, higher dropout rates could degrade model’s performance. For example, a dropout rate of 0.4 results in a sensitivity of only 0.538. This evidence reinforces that excessive regularization hampers the model’s generalization ability, confirming 0.3 as the optimal dropout rate for FCFNet.

## Discussion and Conclusion

In this study, we propose an interpretable factual and counterfactual learning framework (FCFNets) for predicting “at-risk” MASH in young adults with T2D using data from the EHR. Our approach addresses several key challenges in this domain, including cohort imbalance, insufficient precision on the diagnosis of “at-risk” MASH in young adults with T2D and limited prediction interpretability. We leveraged the clinical information of the selected cohorts through data preprocessing and feature selection techniques for the model construction. To balance the dataset while preserving the diversity of the minority class, we employed hybrid oversampling strategies that combine random and dissimilar oversampling methods. Furthermore, by integrating both factual and counterfactual learning, our framework enhanced predictive accuracy and interpretability. The extensive experiments indicate that our proposed FCFNets outperformed all baseline models across different metrics, including sensitivity (0.846), specificity (0.754), accuracy (0.768), and F1-score (0.537). Our model also showed superior counterfactual results by metrics such as validity (1.000) and diversity (0.221). The UNDO oversampling strategy, which effectively balanced random and dissimilar oversampling, was critical to ensure a diverse and unbiased dataset. The SHAP-based global explanations, instance-level interpretations, and feature interaction analyses shed light on the key factors influencing predictions. Notably, features such as “NITRITE UA_result_negative”, “Abnormal results of liver function studies”, and “Vitamin D deficiency” were revealed as significant contributors to be associated with the risk of hepatic fibrosis for young T2D adults. Additionally, counterfactual learning identified important features that could potentially change predictions, such as “Insomnia, unspecified” and “Edema, unspecified”. Furthermore, our feature interaction analysis revealed relationships among variables. For example, the interactions between “NITRITE UA_result_negative” and clinical indicators like “Edema, unspecified” and “Abnormal results of liver function studies” exhibited a strong combined effect on model outcomes.

Despite the promising results, our study has several limitations. First, while leveraging EHR data offers significant predictive power for “at-risk” MASH, it may overlook other relevant data sources, such as imaging or genomic data, that could further enhance model accuracy. Second, the reliance on feature engineering and synthetic oversampling could introduce potential biases for modeling. Third, inherent errors or inconsistencies within EHRs would affect the integrity and completeness of the patient data. In future work, we plan to incorporate other data modalities, such as imaging and proteomics, which can offer a more comprehensive understanding of “at-risk” MASH. We also intend to explore advanced oversampling techniques and more sophisticated counterfactual generation methods to refine model predictions and enhance interpretability. Furthermore, we will deploy our model in real-world clinical settings that allows us to assess its utility and robustness in guiding clinical decision-making for early detection and prevention of “at-risk” MASH in young adults with T2D.

## Supplementary Materials

The codes and supplementary materials are publicly available at: https://github.com/UF-HOBI-Yin-Lab/FCFNets/.

## Data Availability

Data is private, which will not be public available.

## Footnotes

1 https://ufhealth.org/

